# OPEN: A Community-Informed Metadata Framework for Interoperable Computable Phenotypes

**DOI:** 10.64898/2026.11.06.26367456

**Authors:** Jacqueline Honerlaw, Jacqueline A. L. MacArthur, Yuk-Lam Ho, Srushti Gangireddy, Ashley Galloway, Katie Hilty, Gowtham A. Rao, Lang Li, Azza Shoaibi, Spiros Denaxas, Kelly Cho, Wei-Qi Wei

**Affiliations:** VA Boston Healthcare System, Boston, MA, USA; British Heart Foundation Data Science Centre, Health Data Research UK, London, UK; Department of Biomedical Informatics, Vanderbilt University Medical Center, Nashville, TN, USA; Observational Health Data Sciences and Informatics, New York, NY, USA; Department of Biomedical Informatics, College of Medicine, The Ohio State University, Columbus, Ohio, USA; Observational Health Data Analytics, Janssen Research and Development, LLC, Titusville, NJ, USA; Interdisciplinary Transformation University, Linz, Austria; Institute of Health Informatics, University College London, London, UK; Department of Medicine, Mass General Brigham, Harvard Medical School, Boston, MA, USA

**Keywords:** Electronic Health Records, Phenomics, Algorithms, Library, FAIR

## Abstract

**Objective:** To identify community-prioritized metadata for reporting and sharing computable phenotypes (CPs), develop a cross-library candidate framework, assess alignment between user requirements and library metadata, and test the feasibility of metadata exchange.

**Materials and Methods:** We combined an international, library-led effort by the Open PhEnomics Network (OPEN) consortium with a UK community consensus-building exercise aimed at improving the FAIRness (Findable, Accessible, Interoperable, Reusable) of CPs. The consortium performed a cross-library field comparison and literature review to inform candidate fields, refined through consortium consensus. The community approach was developed through stakeholder interviews, a survey, and workshop, guided by a working group. Community requirements were interpretively mapped to the resulting framework and classified into conformance tiers. Interoperability was tested by manually and programmatically mapping diabetes-related phenotypes across libraries.

**Results:** The library-led process produced 23 candidate fields spanning phenotype description, data/technical details, and performance domains. Thirteen fields aligned across all five libraries, and 22 fields aligned with published CP library desiderata. Interoperability testing successfully catalogued phenotypes through manual mapping, with API-mediated mapping achieving 63–76% metadata field completeness across libraries. A community survey (n=124 respondents) identified six metadata elements with majority support as mandatory: description, coding terminology, phenotype type, version history, unique identifier, and data source. Logic representation and validation requirements were rated as mostly context-dependent. Community-identified priorities showed strong alignment with the OPEN framework, though some requirements were only partially represented.

**Discussion:** Through a consensus-based, community-driven approach, we established a CP framework enabling interoperability across libraries.

**Conclusion:** OPEN provides a credible, community-informed foundation for a FAIR CP metadata standard.

## BACKGROUND AND SIGNIFICANCE

Computable phenotypes (CPs) are reproducible algorithms applied to electronic health records (EHR) data to identify diseases, outcomes, exposures, and patient characteristics.[1] They are fundamental for biomedical research, clinical care, population health, and quality improvement.

These algorithms range in complexity from code lists and rule-based logic to artificial intelligence (AI) driven approaches such as natural language processing, machine learning, and large language models. Their development requires substantial clinical expertise, informatics knowledge, terminology mapping, data-model familiarity, and validation effort. Thus, well-documented phenotypes are highly valuable resources for reuse by the broader research community for both observational and interventional research.

Libraries of CPs, also called phenotype repositories, have emerged to support reproducibility and accelerate scientific discovery by cataloging existing algorithms and enabling their reuse. Multiple phenotype libraries have been developed to serve specific communities including common data model users, health systems, and research networks including Phenotype Knowledgebase (PheKB), Centralized Interactive Phenomics Resource (CIPHER), the Health Data Research UK (HDR UK) Phenotype Library, the Observational Health Data Sciences and Informatics (OHDSI) Phenotype Library, and the Vanderbilt Phenotyping Core (VPC) Library.[2-7] The development of community specific phenotype libraries benefits their core users by prioritizing unique needs, such as querying a common data model or accessing control CPs for genetic studies. However, these independently created resources often differ in how they capture, organize, and expose metadata describing phenotype CPs, data sources, logic, validation, performance, provenance, and intended use.

This creates practical barriers. Computable phenotypes are fragmented across libraries, publications and code repositories, and are reported using different terminology and levels of detail. Researchers may struggle to find and compare CPs, judge fitness for purpose, understand validation, or adapt an algorithm to another dataset, coding system, or healthcare setting. Missing links between source, adapted and superseded CPs can lead to duplication, inappropriate reuse, and weak comparability across studies.

The absence of an international standard also means that CPs may lack the metadata needed to be FAIR: findable across distributed resources, accessible in consistent human- and machine-readable forms, interoperable across systems, and reusable with sufficient context.[8] Harmonized metadata are also important for emerging AI-enabled discovery and recommendation, although machine actionability does not require all phenotypes to use one logic format or data model. While previous work has proposed desiderata for phenotype reporting and reuse, no broadly adopted standard defines the core and context-dependent metadata that should accompany shared CPs.[9-17] Furthermore, the creation of such a standard would need to closely align and be informed by the requirements of the stakeholder community using such resources.

Although previous studies have proposed metadata requirements and reporting recommendations for computable phenotypes, a broadly adopted framework supporting interoperability across independently developed phenotype libraries remains lacking. To address this gap, we developed the Open PhEnomics Network (OPEN) framework by harmonizing metadata across five major phenotype libraries and incorporating independently identified community requirements. We further evaluated its practical feasibility through manual and API-based cross-library metadata exchange. Together, these contributions move beyond conceptual recommendations toward an interoperable, community-informed foundation for phenotype discovery, comparison, and reuse.

## OBJECTIVE

To identify community-prioritized metadata for reporting and sharing CPs, develop a cross-library candidate framework, assess alignment between user requirements and library metadata, and test the feasibility of metadata exchange to enable portability.

## MATERIALS AND METHODS

### Study design

We used a convergent design that treated library evidence and community evidence as complementary but distinct contributions to candidate standard development. (Figure 1) The library consortium examined how the absence of a standard had resulted in divergent schemas, developed a common model and tested whether metadata could be exchanged. The community exercise established the consequences of inconsistent reporting and sharing, identified the information users require and explored acceptable approaches to implementation.

**Figure 1.**
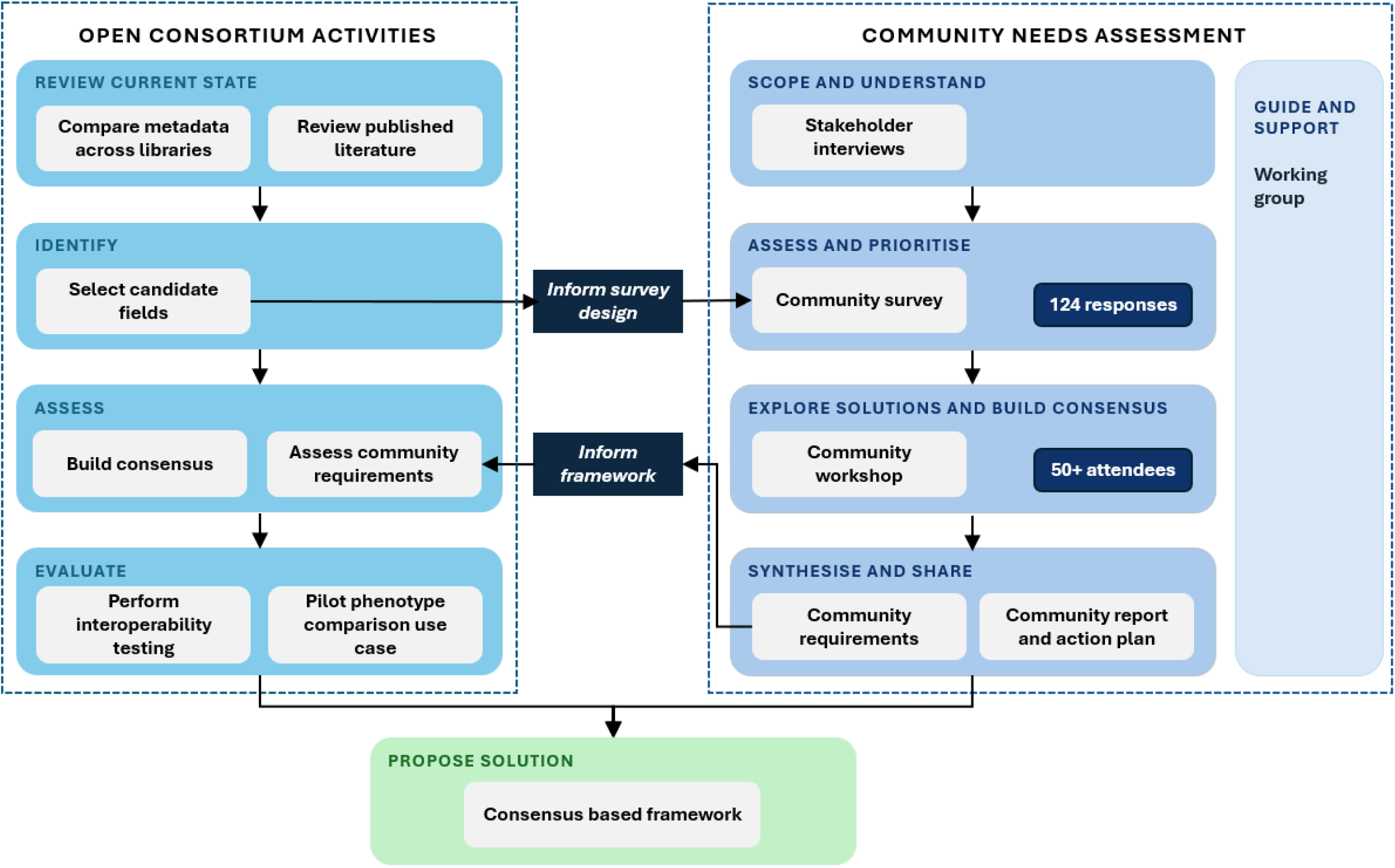
Development of community informed computable phenotype metadata framework. OPEN: Open PhEnomics Network

### International library consortium and scope

The OPEN consortium brought together representatives of PheKB, CIPHER, the OHDSI Phenotype Library, the VPC Library, and HDR UK, together with phenomics experts. The consortium sought to address the metadata divergence that had arisen in the absence of a common reporting and sharing standard by developing a system-agnostic conceptual framework. (Figure 1) Changes to specific library fields or features were out of scope of this effort. Consortium members agreed on six guiding principles to develop the framework. (Table 1)

**Table 1.**
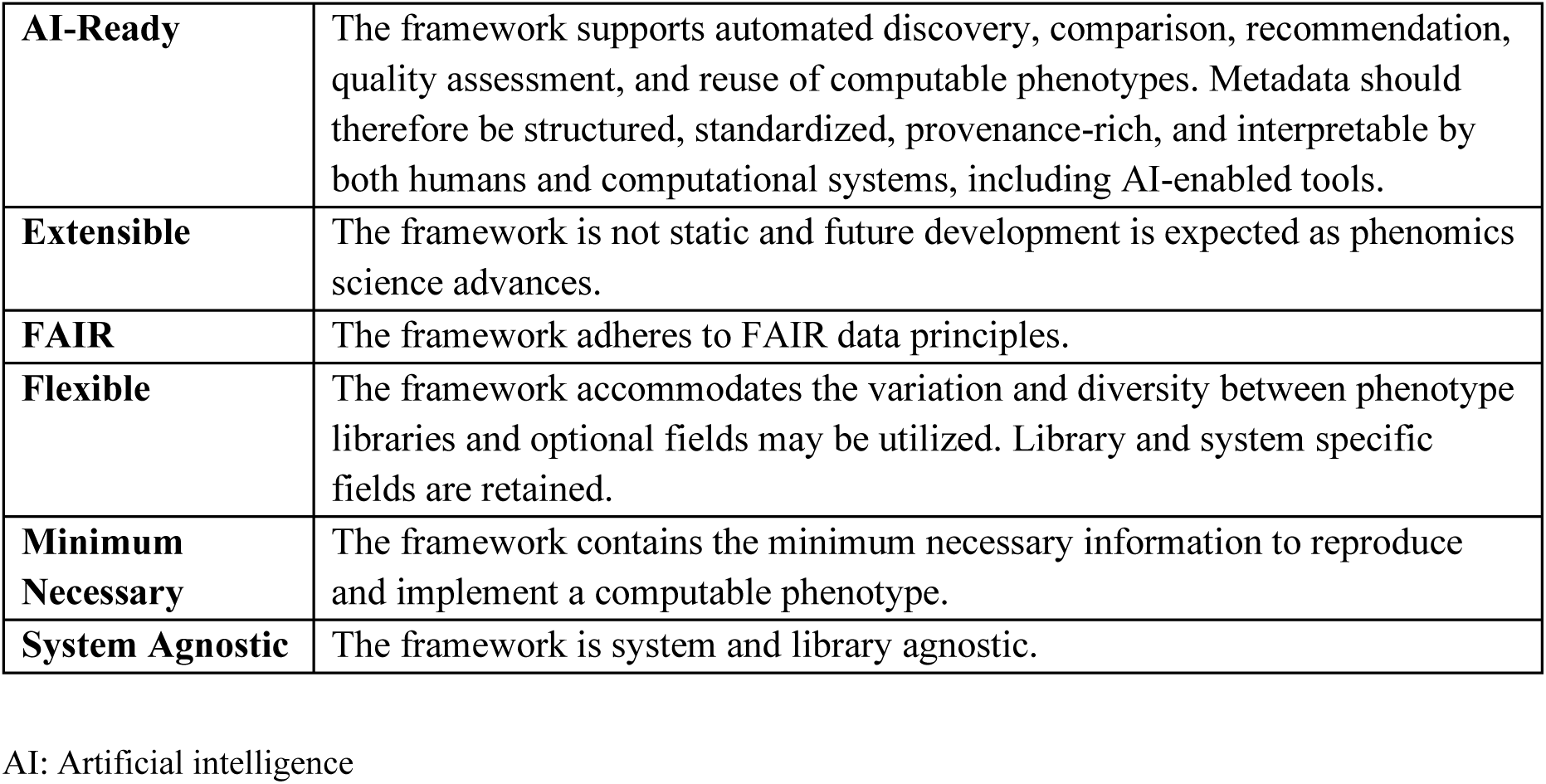
Framework development principles.

#### Cross-library comparison and literature informed candidate field selection

A comparison of the fields available in existing libraries was performed to understand the commonalities between libraries. “Field alignment” indicated that the information is available in the library, though it may not be captured in a structured field and instead may be found in other free text information or attachments. “Partial field alignment” indicated that some information is available, while “field unavailable” means this information is not captured in the library. This effort was built upon earlier work comparing PheKB, HDR UK, and OHDSI phenotype libraries to the CIPHER metadata standard.[16] The comparison was expanded to include the VPC Library.

Candidate fields were informed by the cross-library comparison and by prior literature on phenotype metadata, reporting standards, computable representation, reproducibility, portability, and reuse.[1-17] Literature-derived recommendations were used to identify gaps in existing metadata capture and to assess whether proposed fields were supported by prior work.

#### Consensus building process

The initial set of proposed fields was developed based on library comparison and evaluation of the literature. Library leads had the opportunity to provide feedback on the document and comments were consolidated. Iterative discussions occurred to resolve conflicts, and fields were finalized through a survey.

#### Assessment of community-derived requirements

Community-prioritized metadata concepts and implementation principles were mapped interpretively to OPEN metadata fields. We assessed whether each requirement was directly represented, indirectly represented or absent; whether representation was structured or dependent on free text; and whether the framework supported the user-facing function associated with the requirement. We then classified candidate specification elements into three conformance tiers: Core, Conditional and Extension. This synthesis was not a formal Delphi process and did not alter the historical consortium mapping or survey results.

#### Interoperability testing and pilot use case

We then tested the interoperability of algorithms between libraries after harmonizing them through the framework. We manually mapped a diabetes related, rules-based definition from each source library first to the framework, then to the phenotype model of the destination library, and finally cataloged in the destination library. CIPHER and VPC served as the destination libraries in this test, with VPC, PheKB, and OHDSI phenotypes cataloged in CIPHER, and HDRUK and CIPHER phenotypes cataloged in the VPC library. The mapping was also tested via API with 10 phenotypes from the CIPHER and HDR UK Libraries mapped to the VPC Library. A pilot use case for interoperable phenotypes utilizing the framework was tested with CIPHER’s phenotype comparison tool.

### UK community consensus-building study

An iterative, requirements-driven process was used to identify requirements for FAIR CPs. Stakeholder interviews informed survey design and wider engagement, and a multidisciplinary working group provided expert input and review throughout. (Figure 1) An online survey collected quantitative ratings and free-text responses on current practice, challenges, strategic priorities, metadata and reporting requirements, library functionality and infrastructure needs.

Closed responses were summarized using counts and percentages based on the number answering each question; denominators therefore varied, and questions allowing multiple selections did not sum to 100%. Free-text responses were manually reviewed, summarized, and grouped into recurring themes. Findings were then discussed in a multidisciplinary virtual workshop attended by more than 50 participants.

Workshop sessions tested interpretation of findings, explored areas of agreement and difference, and refined practical priorities. The working group subsequently reviewed the combined evidence and developed recommendations and an action plan. No predefined voting threshold was used; findings are therefore described as community-prioritized or showing broad convergence, rather than formal consensus. Full methods and results are reported in a community report published elsewhere.[18]

## RESULTS

### OPEN phenotype metadata framework and cross-library alignment

The library-led process produced 23 candidate metadata fields across three domains: phenotype description, data and technical details, and performance. (Figure 2) The comparison demonstrated high agreement across libraries, with some evident variation given that they were developed without a common reporting and sharing standard. Thirteen fields aligned across all five libraries, three were absent or only partially represented in one library, and seven showed more variable alignment. Variable fields were designated optional in the framework, including primary use population subcategory, primary use case, URL for source code, and the fields in the performance domain.

**Figure 2.**
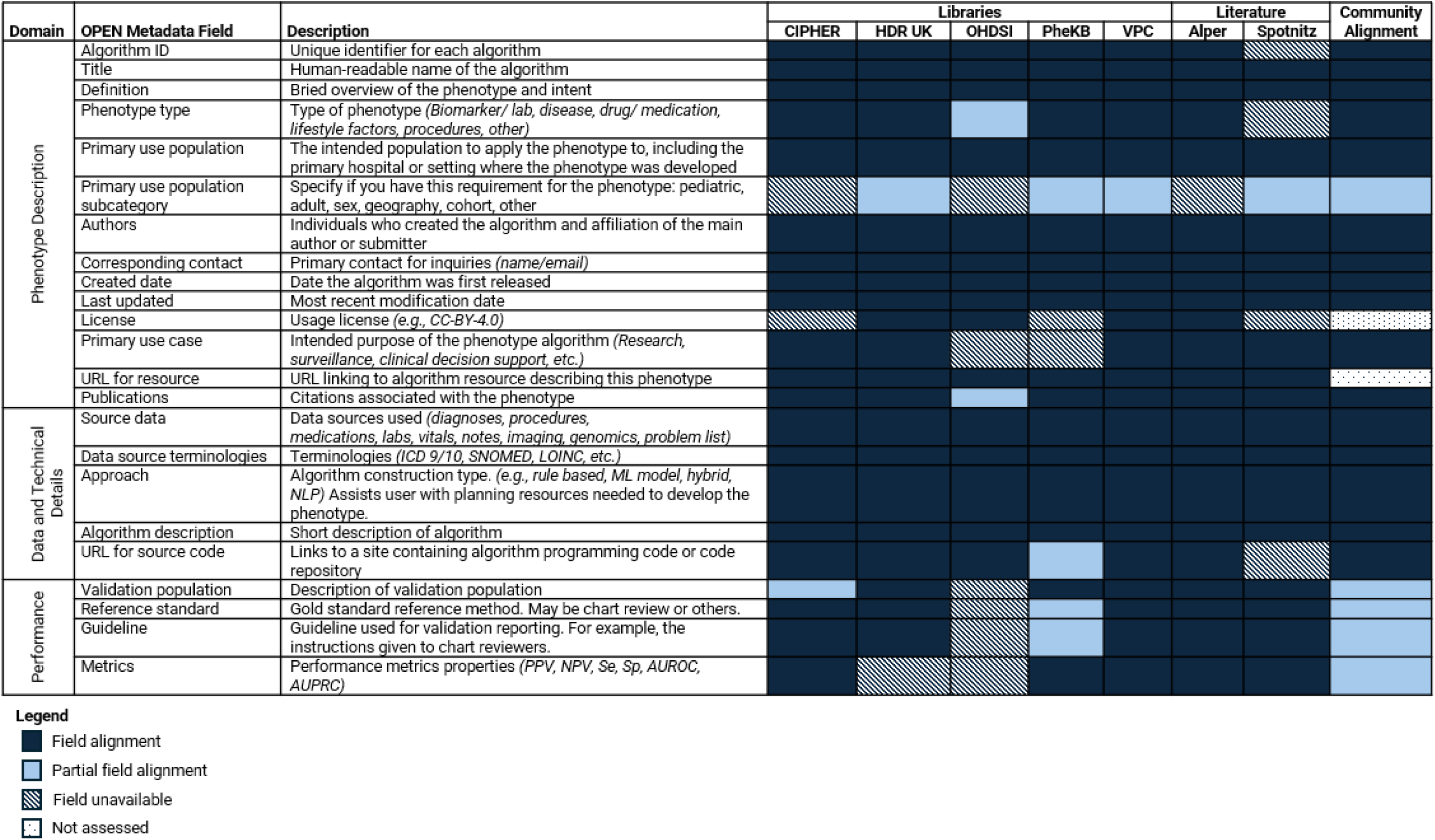
OPEN computable phenotype metadata framework and alignment. AUPRC: Area Under the Precision-Recall Curve; AUROC: Area Under the Receiver Operating Characteristic; CIPHER: Centralized Interactive Phenomics Resource; HDR UK: Health Data Research United Kingdom; ICD: International Classification of Diseases; LOINC: Logical Observation Identifiers Names and Codes; ML: Machine learning; NPV: Negative predictive value; OHDSI: Observational Health Data Sciences and Informatics; OPEN: Open PhEnomics Network; PheKB: Phenotype Knowledgebase; PPV: Positive predictive value; Se: Sensitivity; SNOMED: Systematized Nomenclature of Medicine; Sp: Specificity; VPC: Vanderbilt Phenotyping Core

We also demonstrate the alignment of the OPEN framework against two manuscripts detailing desiderata for phenotype libraries. (Figure 2) Alper and colleagues from the Mobilizing Computable Biomedical Knowledge (MCBK) Community developed categories for metadata capture to make biomedical data FAIR and trustworthy.[12] Twenty-two fields from our framework mapped to elements identified by MCBK which describe the purpose, identification, provenance, technical characteristics, and evidence categories. Spotnitz and colleagues from the Electronic Medical Records and Genomics Network (eMERGE) and OHDSI used a consensus approach to define 39 metadata elements desired for computational phenotypes.[11] Eighteen fields in our metadata framework aligned with their elements.

#### Interoperability testing and pilot use case

The cross-library interoperability test demonstrated successful manual integration of diabetes related phenotype metadata from each library to the CIPHER and VPC libraries using the OPEN framework. (Figure 3, Supplement) The VPC library also integrated 5 phenotypes from CIPHER and 5 from HDR UK into their library using the framework via API. Of the current library schema supported fields from CIPHER and HDR UK, the completeness of the metadata across the sampled phenotypes averaged 76% for HDR UK and 79% for CIPHER. When expanding to view the metadata completeness across all 23 fields of the framework, the populated fields decreased to 59% for HDR and 72% for CIPHER. The API enabled successful mapping of 63% of HDR UK fields and 76% of CIPHER fields into the VPC library.

**Figure 3.**
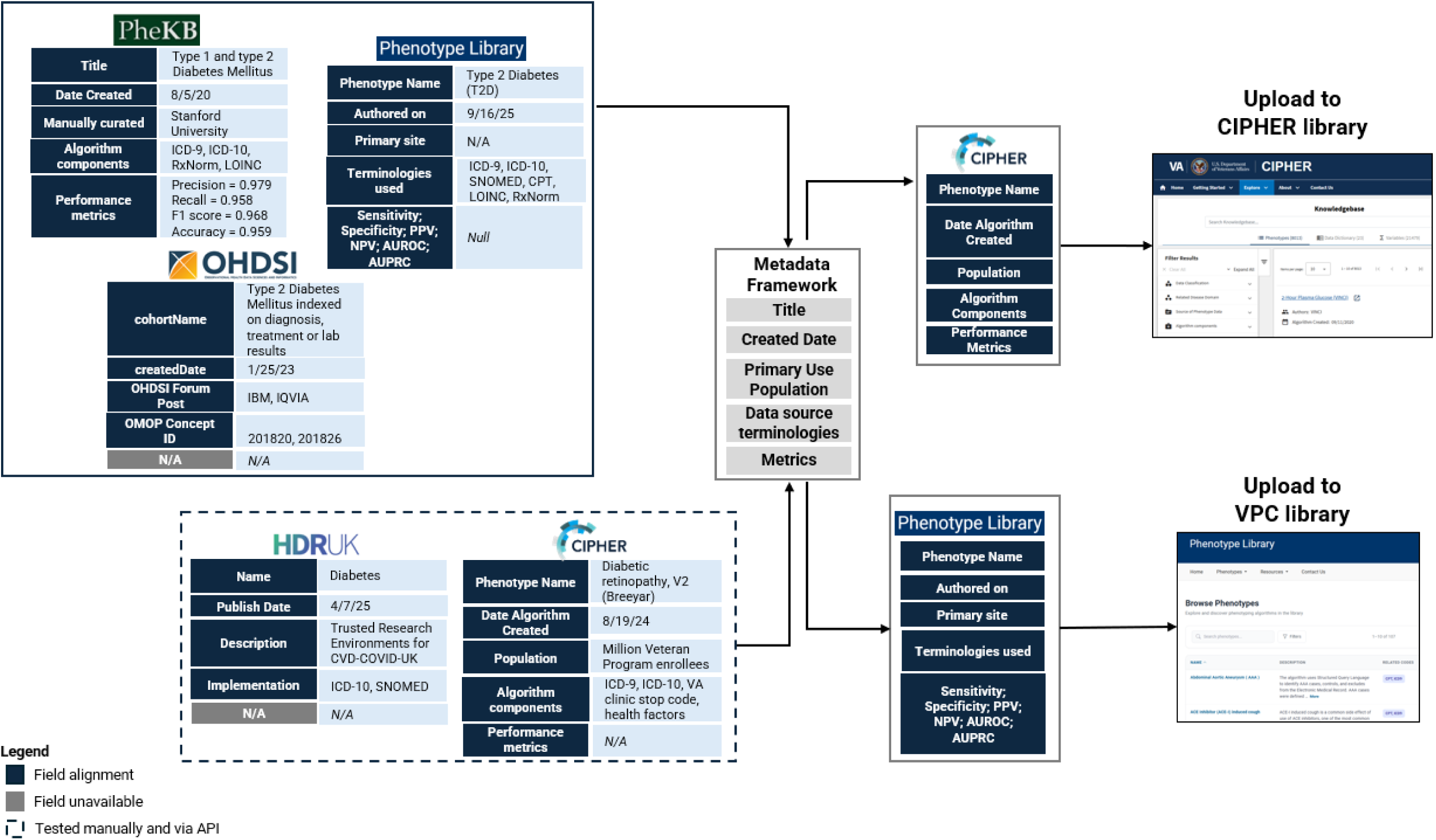
OPEN phenotype metadata framework interoperability testing between libraries. CIPHER: Centralized Interactive Phenomics Resource; CPT: Current Procedural Terminology; HDR UK: Health Data Research United Kingdom; ICD: International Classification of Diseases; LOINC: Logical Observation Identifiers Names and Codes; NPV: Negative predictive value; OHDSI: Observational Health Data Sciences and Informatics; OPEN: Open PhEnomics Network; PheKB: Phenotype Knowledgebase; PPV: Positive predictive value; Se: Sensitivity; SNOMED: Systematized Nomenclature of Medicine; Sp: Specificity; VPC: Vanderbilt Phenotyping Core

CIPHER’s phenotype comparison tool demonstrates the potential impact of an interoperable metadata framework. (Figure 4) If computable phenotype metadata is stored using a common framework, then it can be discoverable and comparable in a central library. This function goes a long way to address community needs regarding consolidated search across libraries, something requested by 75.0% of survey respondents. The phenotype comparison tool is available at <u>phenomics.va.ornl.gov/web/cipher/search</u>.

**Figure 4.**
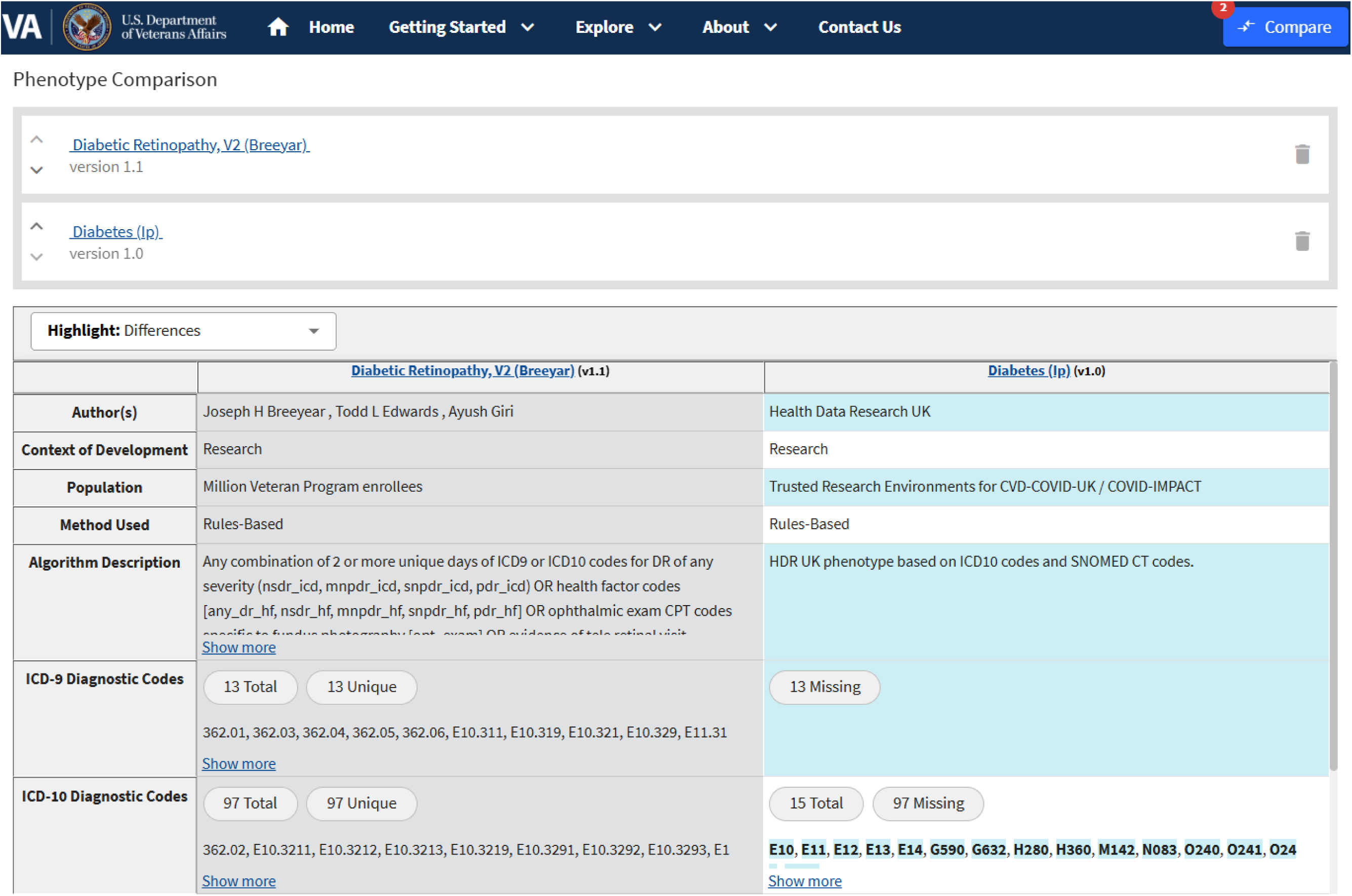
Phenotype comparison use case enabled by consistent metadata capture. Available at phenomics.va.ornl.gov/web/cipher/search.

### UK community consensus-building study

#### Community survey respondent profile, current practices and challenges

The survey received 124 responses, predominantly from UK-affiliated respondents (93.6%) and academia (69.4%). Respondents included researchers (67.7%), data scientists or technical specialists (41.9%), statisticians or methodologists (26.6%), clinicians (12.1%), data custodians or providers (9.7%), and infrastructure or platform providers (8.1%). Respondents could select more than one role.

Respondents worked across the phenotype lifecycle: 79.0% used CPs, 59.7% developed them, 53.2% adapted them, and 30.6% supported relevant infrastructure or tools. CPs were obtained from multiple sources, most commonly publications and phenotype libraries or libraries (both 72.4%), followed by colleagues or collaborations (65.5%), project libraries such as GitHub (44.0%), and trusted research environments or secure data environments (25.9%). Researchers must therefore navigate multiple resources with different formats and levels of documentation.

Challenges were reported throughout the phenotyping lifecycle. Validation (83.7%), adaptation to new datasets or coding systems (82.7%), integration of phenotype-derived information (79.8%), creation (79.0%), and assessment of utility (77.9%) were each rated very or somewhat challenging. Discovery and understanding were each rated very or somewhat challenging by 75.2% of respondents.

#### Community evidence for a coordinated standard and interoperability across libraries

Improved discovery was the clearest infrastructure priority. Respondents selected consolidated search across libraries (75.0%), tools for comparing CPs (69.6%), a standard for sharing across libraries and tools (66.3%), cross-dataset translation tools (63.0%), and validation or benchmarking tools and code-list development tools (both 60.9%).

A standard metadata model for FAIR CPs was the most frequently identified strategic priority (46.7%), followed by improved libraries and discovery tools (45.6%) and tools for translating phenotypes across datasets or coding systems (44.4%). Together with the workshop findings, these results support a community-agreed FAIR standard as a foundation for improving the discovery, assessment, adaptation, and reuse of CPs. Researchers need to find CPs across resources, understand how they differ, and determine their suitability for a particular research question, dataset, population, and setting. Supporting these activities across libraries requires standardized metadata and coordinated infrastructure.

#### Community-prioritized metadata and implementation requirements

Six metadata elements were identified by most survey respondents as mandatory: phenotype description (88.4%), coding terminology (79.6%), phenotype type (77.7%), version history (73.4%), unique identifier (70.2%), and data source or dataset (65.6%) (Table 2). Intended use and phenotyping approach also received strong support, although views differed on whether they should be considered as mandatory fields.

**Table 2.** Community survey results on metadata to accompany shared computable phenotypes.

| **Metadata** | **Mandatory** | **Encouraged** | **Optional** | **Not required** | **Total mandatory + encouraged** |
|---|---|---|---|---|---|
| **Description** | 88% | 7% | 2% | 0% | 96% |
| **Phenotype type e.g. disease, biomarker** | 78% | 13% | 7% | 0% | 90% |
| **Coding terminology** | 80% | 11% | 8% | 0% | 90% |
| **Version history** | 73% | 13% | 9% | 3% | 86% |
| **Unique ID** | 70% | 16% | 2% | 2% | 86% |
| **Data source/dataset** | 66% | 19% | 10% | 2% | 85% |
| **Intended use** | 47% | 31% | 20% | 0% | 78% |
| **Phenotyping approach** | 48% | 27% | 22% | 0% | 75% |
| **Validation approach** | 37% | 37% | 22% | 0% | 73% |
| **Free text description of logic** | 33% | 38% | 25% | 1% | 71% |
| **Publication** | 33% | 37% | 24% | 4% | 70% |
| **Studies/publications used in** | 32% | 37% | 20% | 9% | 69% |
| **Performance metrics** | 28% | 40% | 27% | 1% | 68% |
| **Logic diagram** | 26% | 35% | 33% | 3% | 61% |
| **Link to executable code** | 24% | 37% | 35% | 1% | 60% |

Percentages are based on respondents answering each item and denominators varied between 93 and 95.

Requirements for logic and validation were more context-dependent. Respondents supported providing a representation of phenotype logic but did not identify a single format suitable for all CPs. A standard should therefore specify the information needed to understand and implement a phenotype elsewhere by allowing its logic to be represented through narrative descriptions, diagrams, executable code, or other appropriate formats.

Validation information was strongly valued, but appropriate evidence depended on phenotype type and intended use. A description of the validation approach was rated mandatory by 36.6% and encouraged where relevant by a further 36.6%; performance metrics were rated mandatory by 28.0% and encouraged where relevant by 39.8%. Free-text responses and workshop discussions supported stating whether validation had been undertaken and, where available, reporting the methods, datasets, results, limitations and previous applications.

Respondents cautioned that excessive requirements could discourage sharing. Suggested approaches to reduce burden included structured templates, bulk upload, reusable project-level information, automated metadata capture and progressive levels of completeness. Collectively, these findings support a proportionate model comprising a small core required for all shared CPs, supplemented by context-dependent metadata where applicable. Explicit values such as “not undertaken”, “not applicable”, and “not reported” would allow absent or unavailable evidence to be represented transparently.

### Alignment between community priorities and the OPEN metadata framework

Overall, there was strong alignment between community priorities and the OPEN framework, with most requirements represented in some form (Figure 2). OPEN captured identification, description, phenotype type, coding terminology, source data, approach, intended use, population, and lifecycle dates, together with authorship, licensing, associated publications, algorithm descriptions, source-code links, and validation-related fields.

Some requirements were only partially represented, requiring additional metadata to fully meet community requirements or further work to determine what data should be captured and how it should be structured. (Table 3) This applies to phenotype logic and validation. The community identified a clear need for sufficient information to understand and implement phenotype logic and assess validation, but also recognized that appropriate representations and evidence vary, for example by phenotype type, method, and intended use. The OPEN “Publications” field could similarly be expanded to capture associated publications and uses, distinguishing development, validation, evaluation, and application.

**Table 3.** Alignment between community requirement and OPEN representation, along with assessment and proposed refinement required to establish a community-agreed FAIR standard.

| **Community requirement** | **OPEN representation** | **Assessment and proposed refinement required to meet community requirement** |
|---|---|---|
| Identification and citation | Algorithm ID; resource URL | Strong alignment. Require a globally unique, persistent identifier and an authoritative source record. |
| Description and clinical intent | Title; Definition | Strong alignment. Retain as core metadata and distinguish a concise description from intended clinical meaning where possible. |
| Phenotype type | Phenotype type | Strong alignment. Use an extensible controlled vocabulary. |
| Data and terminology requirements | Data source terminologies; Source data | Strong alignment. Add terminology versions and relevant dataset or common-data-model dependencies. |
| Intended use and population | Primary use case; Primary use population; population subcategory | Represented, but context-dependent. Permit structured categories alongside narrative context. |
| Phenotyping approach | Approach; Algorithm description | Strong alignment. Clarify the method used to construct the phenotype and distinguish it from the executable representation. |
| Logic and implementation | Algorithm description; source-code URL | Partially represented. Record sufficient information to enable the computable phenotype to be reused. Logic representation may vary, but at least one accessible representation of logic should be provided e.g. narrative description, logic diagram, along with source-code. |
| Version history | Created date; Last updated | Partially represented. Add a version identifier, change summary, release status and predecessor/successor relationships. |
| Provenance and relationships between computable phenotypes | Authors; Created date; resource URL | Partially represented. Add links between source, adapted, derived, equivalent, related and superseded computable phenotypes, with the rationale and method for adaptation. |
| Associated publications and uses | Publications | Partially represented. Expand the field to record publications and other persistent links documenting development, validation, adaptation, evaluation or application e.g. clinical trial IDs. Specify the relationship to the phenotype and, where available, the version used, setting and adaptations. |
| Validation evidence and limitations | Validation population; reference standard; guideline; metrics | Partially represented. Add validation status, methods, results, limitations and explicit values for “not undertaken” and “not reported”. Allow for alternative indications of trust and fitness for purpose e.g. clinical adjudication, incidence and prevalence and community endorsement. |
| Maintenance and stewardship | Corresponding contact; Last updated | Partially represented. Add maintainer, review status, maintenance policy and deprecation status. |

Adaptation and relationships between CPs were not explicitly represented in OPEN or consistently captured by participating libraries. Community participants wanted to understand whether a definition had been adapted from another phenotype, how it had changed and how related CPs differed. Representing this information is more complex because it concerns relationships between algorithms rather than attributes of an individual algorithm. Further work is needed to define relationships such as derived from, adapted from, equivalent to, related to, and supersedes.

OPEN therefore provides a strong foundation for a community-informed standard, while identifying areas requiring further discussion and implementation testing. Some gaps may be addressed by refining existing fields; others may require a separate model for relationships between CPs.

## DISCUSSION

We present a community-informed framework for computable phenotype metadata, developed through collaboration among leading phenotype libraries and phenomics experts and assessed against independent evidence from the wider community. The OPEN framework maps heterogeneous library metadata to 23 common fields, enabling CPs to be discovered, assessed and exchanged without requiring libraries to adopt the same internal schema. Manual and API-mediated testing demonstrated the feasibility of transferring metadata between libraries, while the community findings confirmed the need for consistent metadata and coordinated infrastructure to support phenotype discovery, comparison, adaptation, and reuse.

There was strong overall alignment between OPEN and the community requirements, despite the two initiatives approaching the problem from different perspectives. OPEN was informed by current library and curation practices, literature and expert consensus, whereas the community exercise began with user experiences across the phenotype lifecycle. The framework captured the six elements receiving majority support as mandatory: phenotype description, coding terminology, phenotype type, version history, unique identifier, and data source. It also represented intended use, population, phenotyping approach, phenotype logic, associated publications, and validation evidence. This convergence indicates that OPEN captures most of the core information users need to find, understand and assess CPs.

However, agreement on what information is needed does not fully determine how that information should be represented. Although the community work identified clear requirements, the information currently available in libraries does not always meet them. Logic and validation provide two examples of this gap. Sufficient information about phenotype logic is needed to support understanding, implementation and reuse, while validation information should enable users to establish trust and assess fitness for purpose. In both cases, the appropriate content and format will depend on the phenotype type, method, intended use, and evidence available. For example, logic may be represented through narrative descriptions, diagrams, executable code or combinations of these, while relevant validation evidence may include clinical adjudication, community endorsement, comparisons with reference standards, performance measures, or evidence of previous use.

The OPEN framework provides a starting point through algorithm descriptions, source-code links, validation populations, reference standards and performance measures. Where community requirements are only partially represented, further technical development and community input are needed to determine what constitutes sufficient information and how these elements should be structured.

The interoperability results demonstrate both the value and current limitations of a common framework. Manual mapping enabled phenotypes to be catalogued across participating libraries, and API-mediated testing demonstrated the feasibility of automated exchange. However, metadata completeness across all 23 OPEN fields was 59% for HDR UK and 72% for CIPHER, while automated mapping populated 63% and 76% of fields, respectively. A common conceptual framework is therefore necessary but insufficient for complete interoperability while libraries capture different information or represent shared concepts differently. Participating libraries may need to adapt their underlying schemas, while retaining their existing interfaces, specialist fields and distinct roles within the phenomics ecosystem.

The community findings also support a proportionate approach to metadata requirements. A small core can support identification and discovery, while additional information may be required for implementation or assessment of fitness for purpose. Intended use, population, logic representation, and validation evidence may therefore be required when applicable rather than universally mandatory.

Explicit values such as “not undertaken”, “not applicable” and “not reported” could be utilized to distinguish absent evidence from incomplete metadata values. Structured templates, reuse of project-level information, bulk submission and automated metadata capture could reduce contributor burden. Importantly, metadata completeness should not be interpreted as evidence of clinical validity or suitability for every purpose.

Given the infrequent reporting of phenotype metadata in the published literature, we encourage reporting of metadata combined with unique phenotype identifiers in manuscripts and other work products describing computable phenotypes and expansion of journal requirements to capture this information. Recent advances in AI-based text extraction will enable the automated capture of metadata from manuscripts and cataloging in libraries, but the critical first step is for authors to report it.[19, 20]

OPEN is a candidate metadata framework rather than an adopted international standard. The consortium provides technical input from libraries, while the predominantly UK-based community exercise supplies independent evidence of user needs and illustrates strong alignment with the candidate framework. Progression towards a standard will require wider international consultation, stable metadata field definitions, controlled vocabularies where appropriate, and a machine-readable implementation specification defining cardinality, persistent-identifier rules, explicit null values and relationships between metadata fields. Conformance testing should establish whether implementations preserve meaning rather than simply populate similarly named fields. Sustainable governance will also be needed to manage versions, changes, extensions and deprecation.

A major strength of this work is the integration of independent community and technical perspectives, supported by cross-library, API, and cross-system application. We have demonstrated the value of the framework in producing a consolidated search, which could be further improved if the standard was agreed and implemented. The libraries involved in this consortium will work towards a central standard as part of future efforts. Limitations include the self-selected and predominantly academic and UK-affiliated community sample, participation of five libraries, and the limited number of interoperability tests. Field alignment indicated that information was available within a library but did not necessarily mean that it was held in a structured, machine-actionable form. Metadata transfer also did not establish semantic equivalence, clinical validity, transportability or whether implementations identified comparable populations. The framework and proposed refinements therefore require wider consultation and prospective testing.

## CONCLUSION

The OPEN framework provides a consensus-based approach to computable phenotype metadata that supports interoperability, assessment, and scalable reuse across distributed phenotype libraries. Independent community evidence confirms strong alignment with user requirements while identifying areas requiring further development, particularly logic, validation, versioning, associated publications and uses, and relationships between CPs. OPEN provides a credible foundation for a community-informed standard and future AI-enabled applications, but international consultation, technical specification, implementation testing and sustainable governance are required before broader adoption.

## Data Availability

All data produced in the present work are contained in the manuscript.

## ACKNOWLEDGEMENTS

We thank the phenotype library teams, consortium members, working-group members, stakeholder interviewees, survey respondents and workshop participants whose expertise and experience informed this work. Participation in the community exercise or consortium does not necessarily indicate endorsement of every proposed field or conclusion.

This work was supported by the U.S. Department of Veterans Affairs Office of Research and Development. The contents do not represent the views of the U.S. Department of Veterans Affairs or the United States Government.

The UK community work was supported by the British Heart Foundation Data Science Centre (grants SP/19/3/34678 and SI/F/25/21170022), awarded to Health Data Research UK.

## Notes

### Competing Interest Statement

The authors have declared no competing interest.

