## Supplemental tables 1 to 3 for "OPEN: A Community-Informed Metadata Framework for Interoperable Computable Phenotypes"

**Supplementary Table 1.** OPEN phenotype metadata framework

| Domain | Metadata Field | Description | Datatype | Required | Example |
|---|---|---|---|---|---|
| **Phenotype Description** | Algorithm ID | The unique identifier for each algorithm. | String | Yes | Phenotype ID 179 |
| | Title | Human-readable name of the algorithm. | String | Yes | ADHD |
| | Definition | Overview of the phenotype and intent in no more than 3 sentences. | Text | Yes | ICD code and medication based algorithm |
| | Phenotype type | Type of phenotype (e.g., disease, outcome).<br>Properties:<br>biomarker/ lab<br>disease<br>drug/ medication<br>lifestyle factors<br>procedures<br>other | Category | Yes | Disease |
| | Primary use population | The intended population to apply the phenotype to, including the primary hospital or setting where the phenotype was developed. Default all populations, identify subcategory if applicable. | String | Yes | Age > 4; CHOP |
| | Primary use population subcategory | Specify if you have this requirement for the phenotype:<br>pediatric: Yes/no<br>adult: Yes/no<br>sex: Male/Female<br>geography: Free text<br>cohort: Free text<br>other: Free text | String | Optional | Pediatric: Yes |
| | Authors | List of individuals who created the algorithm and affiliation of the main author or submitter. | String | Yes | John Connolly |
| | Corresponding contact | Primary contact (name/email) for inquiries. | String | Yes | John Connolly |
| | Created date | Date the algorithm was first released. | Date | Yes | 9/20/2013 |
| | Last updated | Most recent modification date. | Date | Yes | 9/20/2013 |
| | License | Usage license (e.g., CC-BY-4.0). | String | Optional | *N/A* |

| | | | | | |
|---|---|---|---|---|---|
| | Primary use case | Intended purpose of the phenotype algorithm (research, surveillance, clinical decision support, etc.) | String | Optional | Research |
| | URL for resource | URL linking to algorithm resource. Link out to a general resource describing this phenotype. | URL | Yes | https://phekb.org/phenotype/adhd-phenotype-algorithm |
| | Publications | Citations associated with the phenotype. | String | Yes | *N/A* |
| **Data and Technical Details** | Source data | Which data sources were used?<br>Properties:<br>diagnoses: boolean<br>procedures: boolean<br>medications: boolean<br>labs: boolean<br>vitals: boolean<br>notes: boolean<br>imaging: boolean<br>genomics: boolean<br>problem_list: boolean | Boolean | Yes | Diagnosis: Yes<br>Medications: Yes |
| | Data source terminologies | Terminologies (ICD 9/10, SNOMED, LOINC) | String | Yes | ICD-9; Generic Name, Brand name |
| | Approach | Algorithm construction type (e.g., rule_based, ml_model, hybrid, NLP). Assists user with planning resources needed to develop the phenotype. | Category | Yes | Rule based |
| | Algorithm description | Short description of algorithm. For more information, refer to detailed documentation. | Text | Yes | Cases must be 4 years of age or older, and are defined by a diagnostic history of ADHD/HKD as determined by ICD9 codes and/or a history of ADHD medications… |
| | URL for source code | Links to a site containing algorithm programming code or code repository. | URL | Optional | *N/A* |
| **Performance** | Validation population | Description of validation population. | String | Optional | CHOP |
| | Reference standard | Gold standard reference method. May be chart review or others. | String | Optional | *N/A* |
| | Guideline | Guideline used for validation reporting. For example, the instructions given to chart reviewers. | String | Optional | *N/A* |

| | | | | | |
|---|---|---|---|---|---|
| | Metrics | Performance metrics properties:<br>ppv: float<br>npv: float<br>sensitivity: float<br>specificity: float<br>auroc: float<br>auprc: float | String | Optional | ppv: 0.96 |

Abbreviations: ADHD: Attention-deficit/hyperactivity disorder; AUPRC: Area under the precision-recall curve; AUROC: Area under the receiver operating; CHOP: Children's Hospital of Philadelphia; HKD: Hyperkinetic syndrome; ICD: International Classification of Disease; ID: Identifier; LOINC: Logical Observation Identifiers Names and Codes; NLP: Natural language processing; NPV: Negative predictive value; PPV: Positive predictive value; SNOMED: Systematized Nomenclature of Medicine.

**Supplementary Table 2.** Detailed mapping of community requirements to OPEN fields, including direct, partial and absent representation

| | MetaData Proposal | Description | Datatype | CIPHER | HDR UK | OHDSI | PheKB | VPC | Required Field |
|---|---|---|---|---|---|---|---|---|---|
| Phenotype Description | Algorithm ID | The unique identifier for each algorithm. | string | Yes (Phenotype Identifier) | Yes (phenotype_ID) | Yes (CohortID) | Yes (Phenotype ID) | Yes (Phenotype ID) | **Yes** |
| | Title | Human-readable name of the algorithm. | string | Yes (Phenotype Name) | Yes (name) | Yes (cohortName) | Yes | Yes (Phenotype Name) | **Yes** |
| | Definition | Overview of the phenotype and intent in no more than 3 sentences. | text | Yes (Phenotype Description) | Yes (Description) | Yes (Logic Description) | Yes (Phenotype description) | Yes (Phenotype description) | **Yes** |
| | Phenotype type | Type of phenotype (e.g., disease, outcome).<br>Properties:<br>biomarker/ lab<br>disease<br>drug/ medication<br>lifestyle factors<br>procedures<br>other | category | Yes (Data Classification) | Yes (type) | Partial (clinical description) | Yes (Data source/ clinical domain) | Yes (Phenotype Type) | **Yes** |
| | Primary use population | The intended population to apply the phenotype to, including the primary hospital or setting where the phenotype was developed. Default all populations, identify subcategory if applicable. | string | Yes (Population) | Yes (captured in description) | Yes (OHDSI Forum Post) | Yes | Yes (Primary site) | **Yes** |
| | Primary use population subcategory | Specify if you have this requirement for the phenotype:<br>pediatric: Yes/no<br>adult: Yes/no<br>sex: Male/Female<br>geography: Free text<br>cohort: Free text<br>other: Free text | string | No | Partial (sex) | No | Partial (gender, age) | Partial (sex, age group) | Optional |
| | Authors | List of individuals who created the algorithm and affiliation of the main author or submitter. | string | Yes (Author) | Yes (author) | Yes (contributors, contributor Orc Ids) | Yes (Authors) | Yes (Authors) | **Yes** |

| | | | | | | | | | |
|---|---|---|---|---|---|---|---|---|---|
| | Corresponding contact | Primary contact (name/email) for inquiries. | string | Yes (Contact) | Yes (corresp_author) | Yes (librarian) | Yes | Yes (Contact) | **Yes** |
| | Created date | Date the algorithm was first released. | date | Yes (Date Algorithm Created) | Yes (Publish date) | Yes (createdDate) | Yes (Date Created) | Yes (Authored on) | **Yes** |
| | Last updated | Most recent modification date. | date | Yes (Version Date) | Yes | Yes (Modified Date) | Yes | Yes (Last updated) | **Yes** |
| | License | Usage license (e.g., CC-BY-4.0). | string | No | Yes [Creative Commons Attribution 4.0 (CC-A)] | Yes (Apache License 2.0) | No | Yes (License) | Optional |
| | Primary use case | Intended purpose of the phenotype algorithm (research, surveillance, clinical decision support, etc.) | string | Yes (Context of Development; Role of Phenotype in Analysis) | Yes (captured in description) | No | No | Yes (Primary Use Case) | Optional |
| | URL for resource | URL linking to algorithm resource. Link out to a general resource describing this phenotype. | URL | Yes (Acknowledgement) | Yes | Yes | Yes | Yes (Resource URL) | **Yes** |
| | Publications | Citations associated with the phenotype. | string | Yes (Publication) | Yes (publications) | Partial (OHDSI Forum Post) | Yes | Yes (Publications) | **Yes** |
| **Data and Technical Details** | Source data | Which data sources were used?<br>Properties:<br>diagnoses: boolean<br>procedures: boolean<br>medications: boolean<br>labs: boolean<br>vitals: boolean<br>notes: boolean<br>imaging: boolean<br>genomics: boolean<br>problem_list: boolean | boolean | Yes (Algorithm Components) | Yes (data_sources) | Yes | Yes (Structured data) | Yes (Data source) | **Yes** |
| | Data source terminologies | Terminologies (ICD 9/10, SNOMED, LOINC) | string | Yes (Algorithm Components) | Yes (coding_system) | Yes (OMOP Concept ID) | Yes | Yes (Terminologies used) | **Yes** |
| | Approach | Algorithm construction type (e.g., rule_based, ml_model, hybrid, NLP). Assists user with planning resources | category | Yes (Method Used) | Yes (implementation) | Yes | Yes | Yes (Algorithm type) | **Yes** |

| | | | | | | | | | |
|---|---|---|---|---|---|---|---|---|---|
| | | needed to develop the phenotype. | | | | | | | |
| | Algorithm description | Short description of algorithm. For more information, refer to detailed documentation. | Text | Yes (Algorithm Description) | Yes (within 'implementation ') | Yes (cohort entry event) | Yes | Yes (Algorithm description) | **Yes** |
| | URL for source code | Links to a site containing algorithm programming code or code repository. | URL | Yes (Programming code) | Yes (within 'implementation ') | Yes | Partial | Yes (Source code URL) | Optional |
| **Performance** | Validation population | Description of validation population. | string | Partial (Description of Validation) | Yes (validation) | No | Yes | Yes (Validation population) | Optional |
| | Reference standard | Gold standard reference method. May be chart review or others. | string | Yes (Description of Validation) | Yes (either within 'validation' or endorsements) | No | Partial - may be in attachment | Yes (Reference standard) | Optional |
| | Guideline | Guideline used for validation reporting. For example, the instructions given to chart reviewers. | string | Yes(Description of Validation) | Yes (within 'validation') | No | Partial - may be in attachment | Yes (Guideline) | Optional |
| | Metrics | Performance metrics properties:<br>ppv: float<br>npv: float<br>sensitivity: float<br>specificity: float<br>auroc: float<br>auprc: float | string | Yes (Performance Metrics) | No | No | Yes | Yes (Performance Metrics Sensitivity; Specificity; PPV; NPV; AUROC; AUPRC) | Optional |

**Supplementary Table 3.** Phenotypes used in library interoperability testing

| Source Library | Phenotype Name | Source Library URL | Mapping Method | Destination Library |
|---|---|---|---|---|
| HDR UK | Diabetes | https://phenotypes.healthdatagateway.org/phenotypes/PH1741/detail/ | Manual & API | VPC |
| HDR UK | COVID-19 infection | https://phenotypes.healthdatagateway.org/phenotypes/PH1/detail/ | Manual & API | VPC |
| HDR UK | Asthma | https://phenotypes.healthdatagateway.org/phenotypes/PH12/detail/ | Manual & API | VPC |
| HDR UK | Chronic Kidney Disease (CKD) | https://phenotypes.healthdatagateway.org/phenotypes/PH22/detail/ | Manual & API | VPC |
| HDR UK | Secondary pulmonary hypertension | https://phenotypes.healthdatagateway.org/phenotypes/PH4349/detail/ | Manual & API | VPC |
| CIPHER | Acute Myocardial Infarction (Bray) | https://phenomics.va.ornl.gov/web/cipher/phenotype-viewer/details?uqid=0643b91dfd5a4926ac4592463f02a30b&name=Acute_Myocardial_Infarction__Bray_ | Manual & API | VPC |
| CIPHER | Weight loss (PERC) | https://phenomics.va.ornl.gov/web/cipher/phenotype-viewer/details?uqid=bc019b69d46248d9a632d6dc6ae16adb&name=Weight_loss__PERC_ | Manual & API | VPC |
| CIPHER | Diabetic Retinopathy, V2 (Breeyar) | https://phenomics.va.ornl.gov/web/cipher/phenotype-viewer/details?uqid=7c69b02c5d7d4a699b6cb2f90ccb3eaf&name=Diabetic_Retinopathy__V2__Breeyar_ | Manual & API | VPC |
| CIPHER | Alzheimer's disease (MAP) | https://phenomics.va.ornl.gov/web/cipher/phenotype-viewer/details?uqid=e9c7b53445af46f6b2ad8ba334d5288d&name=Alzheimer_s_disease__MAP_ | Manual & API | VPC |
| CIPHER | Coronary atherosclerosis (MAP) | https://phenomics.va.ornl.gov/web/cipher/phenotype-viewer/details?uqid=eadcd630ab544709eab0b29e6e1a2dfc&name=Coronary_atherosclerosis__MAP_ | Manual & API | VPC |
| VPC | Type 2 Diabetes | https://phenotyping.org/phenotype-detail/93 | Manual | CIPHER |
| PheKB | Type 1 and type 2 Diabetes Mellitus | https://phekb.org/phenotype/type-1-and-type-2-diabetes-mellitus | Manual | CIPHER |
| OHDSI | Type 2 Diabetes Mellitus indexed on diagnosis, treatment or lab results | https://github.com/OHDSI/PhenotypeLibrary/blob/main/inst/sql/sql_server/288.sql | Manual | CIPHER |
