## Supplemental table 4 for "OPEN: A Community-Informed Metadata Framework for Interoperable Computable Phenotypes"

**Supplementary Table 4: CIPHER to VPC Library Mapping Results Summary**

| Metadata Framework Field | In CIPHER Schema (Yes/ No) | Acute Myocardial Infarction (Bray) | Weight loss (PERC) | Diabetic Retinopathy, V2 (Breeyar) | Alzheimer's disease (MAP) | Coronary atherosclerosis (MAP) | CIPHER Field Completion | In VPC Schema (Yes/ No) | Acute Myocardial Infarction (Bray) | Weight loss (PERC) | Diabetic Retinopathy, V2 (Breeyar) | Alzheimer's disease (MAP) | Coronary atherosclerosis (MAP) | VPC Field Completion |
|---|---|---|---|---|---|---|---|---|---|---|---|---|---|---|
| Algorithm ID | Y | 1 | 1 | 1 | 1 | 1 | **1** | Y | 1 | 1 | 1 | 1 | 1 | **1** |
| Title | Y | 1 | 1 | 1 | 1 | 1 | **1** | Y | 1 | 1 | 1 | 1 | 1 | **1** |
| Definition | Y | 1 | 1 | 1 | 1 | 1 | **1** | Y | 1 | 1 | 1 | 1 | 1 | **1** |
| Phenotype type | Y | 1 | 1 | 1 | 1 | 1 | **1** | Y | 1 | 1 | 1 | 1 | 1 | **0.9** |
| Primary use population | Y | 0 | 1 | 1 | 1 | 1 | **0.8** | Y | 0 | 1 | 1 | 1 | 1 | **0.4** |
| Primary use population subcategory | N | *N/A* | *N/A* | *N/A* | *N/A* | *N/A* | **N/A** | Y | 0 | 0 | 0 | 0 | 0 | **0.5** |
| Authors | Y | 1 | 1 | 1 | 1 | 1 | **1** | Y | 1 | 1 | 1 | 1 | 1 | **1** |
| Corresponding contact | Y | 1 | 1 | 1 | 1 | 1 | **1** | Y | 1 | 1 | 1 | 1 | 1 | **1** |
| Created date | Y | 1 | 1 | 1 | 1 | 1 | **1** | Y | 1 | 1 | 1 | 1 | 1 | **0.9** |
| Last updated | Y | 1 | 1 | 1 | 0 | 1 | **0.8** | Y | 1 | 1 | 1 | 0 | 1 | **0.4** |
| License | N | *N/A* | *N/A* | *N/A* | *N/A* | *N/A* | **N/A** | Y | 0 | 0 | 0 | 0 | 0 | **0.5** |
| Primary use case | Y | 1 | 1 | 1 | 1 | 1 | **1** | Y | 1 | 1 | 1 | 1 | 1 | **1** |
| URL for resource | Y | *N/A* | 0 | 0 | 0 | 0 | **0** | Y | 1 | 1 | 1 | 1 | 1 | **0.9** |
| Publications | Y | 1 | 0 | 1 | 1 | 1 | **0.8** | Y | 1 | 0 | 1 | 1 | 1 | **0.8** |
| Source data | Y | 0 | 1 | 1 | 1 | 1 | **0.8** | Y | 0 | 1 | 1 | 1 | 1 | **0.9** |
| Data source terminologies | Y | 1 | 1 | 1 | 1 | 1 | **1** | Y | 1 | 1 | 1 | 1 | 1 | **1** |
| Approach | Y | 1 | 1 | 1 | 1 | 1 | **1** | Y | 1 | 1 | 1 | 1 | 1 | **1** |
| Algorithm description | Y | 1 | 1 | 1 | 1 | 1 | **1** | Y | 1 | 1 | 1 | 1 | 1 | **0.5** |
| URL for source code | Y | 0 | 0 | 0 | 0 | 0 | **0** | Y | 0 | 0 | 0 | 0 | 0 | **0.3** |
| Validation population | Y | 0 | 1 | 1 | 1 | 1 | **0.8** | Y | 0 | 1 | 0 | 1 | 1 | **0.6** |
| Reference standard | Y | 0 | 1 | 0 | 1 | 1 | **0.6** | Y | 0 | 1 | 0 | 1 | 1 | **0.6** |
| Guideline | Y | 0 | 1 | 0 | 1 | 1 | **0.6** | Y | 0 | 1 | 0 | 1 | 1 | **0.5** |
| Metrics | Y | 0 | 0 | 0 | 1 | 1 | **0.4** | Y | 0 | 0 | 0 | 1 | 1 | **0.4** |
| Fields populated of the 23 framework fields | | 13 | 17 | 16 | 18 | 19 | | | 14 | 18 | 16 | 19 | 20 | |
| Completion, all 23 fields | | 57% | 74% | 70% | 78% | 83% | | | 61% | 78% | 70% | 83% | 87% | |
| Completion, schema-supported fields | | 62% | 81% | 76% | 86% | 90% | | | 61% | 78% | 70% | 83% | 87% | |
| Library mean, all 23 fields | 72% | | | | | | | 76% | | | | | | |
| Library mean, schema-supported fields only | 79% | | | | | | | 76% | | | | | | |
