## Supplemental table 5 for "OPEN: A Community-Informed Metadata Framework for Interoperable Computable Phenotypes"

**Supplementary Table 5: CIPHER to VPC Library Mapping**

| CIPHER Field | CIPHER Value | Field Complete (0/1) | Metadata Framework Field | VPC Library Field | VPC Library Value | Field Complete (0/1) | Note |
|---|---|---|---|---|---|---|---|
| Phenotype ID | 0643b91dfd5a4926ac4592463f02a30b | 1 | Algorithm ID | Phenotype ID | 105 | 1 | |
| Phenotype Name | Acute Myocardial Infarction (Bray) | 1 | Title | Phenotype Name | Acute Myocardial Infarction (Bray) | 1 | |
| Phenotype Description | HDR UK phenotype based on ICD10 codes ar | 1 | Definition | Phenotype Description | HDR UK phenotype based on ICD10 codes and SNOMED CT code | 1 | |
| Data Classification | Diseases | 1 | Phenotype type | Phenotype Type | Disease or Syndrome | 1 | |
| Population | *Null* | 0 | Primary use population | Primary Site | *Null* | 0 | |
| *N/A* | *N/A* | *N/A* | Primary use population subcategory | Sex; Age Group; Age Range; Race; Ancestry; | | 0 | |
| Author(s) | Health Data Research UK | 1 | Authors | Authors | Health Data Research UK | 1 | |
| Contact | CIPHER | 1 | Corresponding contact | Contact | | 1 | |
| Date Algorithm Created | 5/7/2025 | 1 | Created date | Authored on | 7-May-25 | 1 | |
| Version Date | 16-Apr-26 | 1 | Last updated | Last Updated | 20-Aug-26 | 1 | |
| *N/A* | *N/A* | *N/A* | License | License | | 0 | |
| Context of Development | Research | 1 | Primary use case | Primary Use Case | Research | 1 | |
| Acknowledgement | *Null* | *N/A* | URL for resource | Resource URL | | 1 | CIPHER URL |
| Publication | https://phenotypes.healthdatagateway.org/ | 1 | Publications | Publications | https://phenotypes.healthdatagateway.org/phenotypes/PH179( | 1 | |
| Source of phenotype data | *Null* | 0 | Source data | Data Source | no data | 0 | |
| Algorithm components | ICD-10, SNOMED | 1 | Data source terminologies | Terminologies Used | ICD-10, SNOMED CT | 1 | |
| Method used | Rules-Based (i.e., only structured data were | 1 | Approach | Algorithm Type | Rule-based | 1 | |
| Algorithm description | HDR UK phenotype based on ICD10 codes ar | 1 | Algorithm description | Algorithm Description | HDR UK phenotype based on ICD10 codes and SNOMED CT code | 1 | |
| Programming code | *Null* | 0 | URL for source code | Source Code URL | | 0 | |
| Description of Validation | *Null* | 0 | Validation population | Validation Population | | 0 | |
| Description of Validation | *Null* | 0 | Reference standard | Reference Standard | | 0 | |
| Description of Validation | *Null* | 0 | Guideline | Guideline | | 0 | |
| Performance metrics | *Null* | 0 | Metrics | Performance Metrics Sensitivity; Specificity; PPV; NPV; AUROC; AUPRC |  | 0 |  |

| CIPHER Field | CIPHER Value | Field Complete (0/1) | Metadata Framework Field | VPC Library Field | VPC Library Value | Field Complete (0/1) | Note |
|---|---|---|---|---|---|---|---|
| Phenotype ID | bc019b69d46248d9a632d6dc6ae16adb | 1 | Algorithm ID | Phenotype ID | 106 | 1 | |
| Phenotype Name | Weight loss (PERC) | 1 | Title | Phenotype Name | Weight loss (PERC) | 1 | |
| Phenotype Description | Elixhauser set initially based on Quan H 2005 | 1 | Definition | Phenotype Description | Elixhauser set initially based on Quan H 2005, but now based on | 1 | |
| Data Classification | Diseases | 1 | Phenotype type | Phenotype Type | Disease or Syndrome | 1 | |
| Population | All active VHA users (at least 1 appointment | 1 | Primary use population | Primary Site | All active VHA users (at least 1 appointment or hospital stay) wit | 1 | |
| *N/A* | *N/A* | *N/A* | Primary use population subcategory | Sex; Age Group; Age Range; Race; Ancestry; | | 0 | |
| Author(s) | Program Evaluation and Resource Center, Office of Mental Health and Suicide Prevention. | 1 | Authors | Authors | Program Evaluation and Resource Center, Office of Mental Health and Suicide Prevention. | 1 | |
| Contact | CIPHER | 1 | Corresponding contact | Contact | | 1 | |
| Date Algorithm Created | 10/1/2015 | 1 | Created date | Authored on | 10-Jan-15 | 1 | |
| Version Date | Mar 25, 2025 | 1 | Last updated | Last Updated | Mar 25, 2025 | 1 | |
| *N/A* | *N/A* | *N/A* | License | License | | 0 | |
| Context of Development | Healthcare Operations | 1 | Primary use case | Primary Use Case | Healthcare Operations | 1 | |
| Acknowledgement | *Null* | 0 | URL for resource | Resource URL | | 1 | CIPHER URL |
| Publication | *Null* | 0 | Publications | Publications | | 0 | |
| Source of phenotype data | CDW (Corporate Data Warehouse) | 1 | Source data | Data Source | CDW (Corporate Data Warehouse) | 1 | |
| Algorithm components | ICD-10 | 1 | Data source terminologies | Terminologies Used | ICD-10 | 1 | |
| Method used | Rules-Based (i.e., only structured data were | 1 | Approach | Algorithm Type | Rule-based | 1 | |
| Algorithm description | At least one encounter or inpatient stay witl | 1 | Algorithm description | Algorithm Description | At least one encounter or inpatient stay with the diagnosis withi | 1 | |
| Programming code | *Null* | 0 | URL for source code | Source Code URL | | 0 | |
| Description of Validation | All active VHA users (at least 1 appointment | 1 | Validation population | Validation Population | All active VHA users (at least 1 appointment or hospital stay) wit | 1 | |
| Description of Validation | Validated by subject matter experts and clin | 1 | Reference standard | Reference Standard | Validated by subject matter experts and clinical field staff. | 1 | |
| Description of Validation | Validated by subject matter experts and clin | 1 | Guideline | Guideline | Validated by subject matter experts and clinical field staff. | 1 | |
| Performance metrics | *Null* | 0 | Metrics | Performance Metrics Sensitivity; Specificity; PPV; NPV; AUROC; AUPRC |  | 0 |  |

| CIPHER Field | CIPHER Value | Field Complete (0/1) | Metadata Framework Field | VPC Library Field | VPC Library Value | Field Complete (0/1) | Note |
|---|---|---|---|---|---|---|---|
| Phenotype ID | 7c69b02c5d7d4a699b6cb2f90ccb3eaf | 1 | Algorithm ID | Phenotype ID | 107 | 1 | |
| Phenotype Name | Diabetic Retinopathy, V2 (Breeyar) | 1 | Title | Phenotype Name | Diabetic Retinopathy, V2 (Breeyar) | 1 | |
| Phenotype Description | Find indivdiuals with verifiable evidence of d | 1 | Definition | Phenotype Description | Find indivdiuals with verifiable evidence of diabetic retinopathy | 1 | |

| | | | | | | | |
|---|---|---|---|---|---|---|---|
| Data Classification | Diseases | 1 | Phenotype type | Phenotype Type | Disease or Syndrome | 1 | |
| Population | Million Veteran Program enrollees | 1 | Primary use population | Primary Site | Million Veteran Program enrollees | 1 | |
| *N/A* | *N/A* | *N/A* | Primary use population subcategory | Sex; Age Group; Age Range; Race; Ancestry; | | 0 | |
| Author(s) | Joseph H Breeyear, Todd L Edwards, Ayush Giri | 1 | Authors | Authors | Joseph H Breeyear, Todd L Edwards, Ayush Giri | 1 | |
| Contact | | 1 | Corresponding contact | Contact | | 1 | |
| Date Algorithm Created | 8/19/2024 | 1 | Created date | Authored on | 19-Aug-24 | 1 | |
| Version Date | 12-Aug-26 | 1 | Last updated | Last Updated | 12-Aug-26 | 1 | |
| *N/A* | *N/A* | *N/A* | License | License | | 0 | |
| Context of Development | Research | 1 | Primary use case | Primary Use Case | Research | 1 | |
| Acknowledgement | *Null* | 0 | URL for resource | Resource URL | | 1 | CIPHER URL |
| Publication | https://pubmed.ncbi.nlm.nih.gov/39158361, | 1 | Publications | Publications | https://pubmed.ncbi.nlm.nih.gov/39158361/ | 1 | |
| Source of phenotype data | MVP (Million Veteran Program) | 1 | Source data | Data Source | MVP (Million Veteran Program) | 1 | |
| Algorithm components | ICD9, ICD10, VA Clinic Stop Codes, Other Hea | 1 | Data source terminologies | Terminologies Used | ICD9, ICD10, VA Clinic Stop Codes, Other Health Factor Codes | 1 | |
| Method used | Rules-Based (i.e., only structured data were | 1 | Approach | Algorithm Type | Rule-based | 1 | |
| Algorithm description | Any combination of 2 or more unique days o | 1 | Algorithm description | Algorithm Description | Any combination of 2 or more unique days of ICD9 or ICD10 cod | 1 | |
| Programming code | *Null* | 0 | URL for source code | Source Code URL | | 0 | |
| Description of Validation | *Null* | 1 | Validation population | Validation Population | | 0 | |
| Description of Validation | *Null* | 0 | Reference standard | Reference Standard | | 0 | |
| Description of Validation | *Null* | 0 | Guideline | Guideline | | 0 | |
| Performance metrics | *Null* | 0 | Metrics | Performance Metrics Sensitivity; Specificity; PPV; NPV; AUROC; AUPRC | | 0 | |

| **CIPHER Field** | **CIPHER Value** | **Field Complete (0/1)** | **Metadata Framework Field** | **VPC Library Field** | **VPC Library Value** | **Field Complete (0/1)** | **Note** |
|---|---|---|---|---|---|---|---|
| Phenotype ID | e9c7b53445af46f6b2ad8ba334d5288d | 1 | Algorithm ID | Phenotype ID | 108 | 1 | |
| Phenotype Name | Alzheimer's disease (MAP) | 1 | Title | Phenotype Name | Alzheimer's disease (MAP) | 1 | |
| Phenotype Description | Condition defined using MAP unsupervised c | 1 | Definition | Phenotype Description | Condition defined using MAP unsupervised clustering algorithm. | 1 | |
| Data Classification | Diseases | 1 | Phenotype type | Phenotype Type | Disease or Syndrome | 1 | |
| Population | MVP | 1 | Primary use population | Primary Site | MVP | 1 | |
| *N/A* | *N/A* | *N/A* | Primary use population subcategory | Sex; Age Group; Age Range; Race; Ancestry; | | 0 | |
| Author(s) | MVP Core | 1 | Authors | Authors | MVP Core | 1 | |
| Contact | | 1 | Corresponding contact | Contact | | 1 | |
| Date Algorithm Created | 11/1/2019 | 1 | Created date | Authored on | 1-Nov-19 | 1 | |
| Version Date | *Null* | 0 | Last updated | Last Updated | | 0 | |
| *N/A* | *N/A* | *N/A* | License | License | | 0 | |
| Context of Development | Research | 1 | Primary use case | Primary Use Case | Research | 1 | |
| Acknowledgement | *Null* | 0 | URL for resource | Resource URL | | 1 | CIPHER URL |
| Publication | https://www.ncbi.nlm.nih.gov/pubmed/316 | 1 | Publications | Publications | https://www.ncbi.nlm.nih.gov/pubmed/31613361 | 1 | |
| Source of phenotype data | CDW (Corporate Data Warehouse) | 1 | Source data | Data Source | CDW (Corporate Data Warehouse) | 1 | |
| Algorithm components | ICD9, ICD10, CUI | 1 | Data source terminologies | Terminologies Used | ICD9, ICD10, Text Snippets CUI | 1 | |
| Method used | Machine learning: Unsupervised | 1 | Approach | Algorithm Type | Machine learning Model | 1 | |
| Algorithm description | MAP is an unsupervised clustering algorithm | 1 | Algorithm description | Algorithm Description | MAP is an unsupervised clustering algorithm that uses counts of | 1 | |
| Programming code | *Null* | 0 | URL for source code | Source Code URL | | 0 | |
| Description of Validation | MVP | 1 | Validation population | Validation Population | MVP | 1 | |
| Description of Validation | A total of 100 patient charts were sampled f | 1 | Reference standard | Reference Standard | A total of 100 patient charts were sampled from the MVP cohort | 1 | |
| Description of Validation | A total of 100 patient charts were sampled f | 1 | Guideline | Guideline | A total of 100 patient charts were sampled from the MVP cohort | 1 | |
| Performance metrics | Sensitivity: 0.35; NPV: 0.83; Specificity: 1; PP | 1 | Metrics | Performance Metrics Sensitivity; Specificity; PPV; NPV; AUROC; AUPRC | Sensitivity: 0.35; NPV: 0.83; Specificity: 1; PPV: 1 | 1 | |

| **CIPHER Field** | **CIPHER Value** | **Field Complete (0/1)** | **Metadata Framework Field** | **VPC Library Field** | **VPC Library Value** | **Field Complete (0/1)** | **Note** |
|---|---|---|---|---|---|---|---|
| Phenotype ID | eadcd630ab544709eab0b29e6e1a2dfc | 1 | Algorithm ID | Phenotype ID | 109 | 1 | |
| Phenotype Name | Coronary atherosclerosis (MAP) | 1 | Title | Phenotype Name | Coronary atherosclerosis (MAP) | 1 | |
| Phenotype Description | Condition defined using MAP unsupervised c | 1 | Definition | Phenotype Description | Condition defined using MAP unsupervised clustering algorithm. | 1 | |
| Data Classification | Diseases | 1 | Phenotype type | Phenotype Type | Disease or Syndrome | 1 | |
| Population | MVP | 1 | Primary use population | Primary Site | MVP | 1 | |
| *N/A* | *N/A* | *N/A* | Primary use population subcategory | Sex; Age Group; Age Range; Race; Ancestry; | | 0 | |
| Author(s) | MVP Core | 1 | Authors | Authors | MVP Core | 1 | |
| Contact | | 1 | Corresponding contact | Contact | | 1 | |
| Date Algorithm Created | 11/1/2019 | 1 | Created date | Authored on | 1-Nov-19 | 1 | |
| Version Date | 10-Dec-24 | 1 | Last updated | Last Updated | 10-Dec-24 | 1 | |

| | | | | | | |
|---|---|---|---|---|---|---|---|
| *N/A* | *N/A* | *N/A* | License | License | | 0 | |
| Context of Development | Research | 1 | Primary use case | Primary Use Case | Research | 1 | |
| Acknowledgement | *Null* | 0 | URL for resource | Resource URL | | 1 | CIPHER URL |
| Publication | https://www.ncbi.nlm.nih.gov/pubmed/316 | 1 | Publications | Publications | https://www.ncbi.nlm.nih.gov/pubmed/31613361 | 1 | |
| Source of phenotype data | CDW (Corporate Data Warehouse) | 1 | Source data | Data Source | CDW (Corporate Data Warehouse) | 1 | |
| Algorithm components | ICD9, ICD10, Text Snippets CUI | 1 | Data source terminologies | Terminologies Used | ICD9, ICD10, Text Snippets CUI | 1 | |
| Method used | Machine learning: Unsupervised | 1 | Approach | Algorithm Type | Machine Learning | 1 | |
| Algorithm description | MAP is an unsupervised clustering algorithm | 1 | Algorithm description | Algorithm Description | MAP is an unsupervised clustering algorithm that uses counts of | 1 | |
| Programming code | *Null* | 0 | URL for source code | Source Code URL | | 0 | |
| Description of Validation | MVP | 1 | Validation population | Validation Population | MVP | 1 | |
| Description of Validation | A total of 200 patient charts were sampled f | 1 | Reference standard | Reference Standard | A total of 200 patient charts were sampled from the MVP cohort | 1 | |
| Description of Validation | A total of 200 patient charts were sampled f | 1 | Guideline | Guideline | A total of 200 patient charts were sampled from the MVP cohort | 1 | |
| Performance metrics | FPR: 0.05; Sensitivity: 0.107; NPV: 0.298; AU | 1 | Metrics | Performance Metrics Sensitivity; Specificity; PPV; NPV; AUROC; AUPRC | Sensitivity: 0.107; NPV: 0.298; AUC: 0.675; PPV: 0.842 | 1 | |
