## Supplemental table 6 for "OPEN: A Community-Informed Metadata Framework for Interoperable Computable Phenotypes"

**Supplementary Table 6: HDR UK to VPC Library Mapping Results Summary**

| Metadata Framework Field | In HDR UK Schema (Yes/ No) | Diabetes | COVID-19 infection | Asthma | Chronic Kidney Disease (CKD) | Secondary pulmonary hypertension | HDR UK Field Completion | In VPC Schema (Yes/ No) | Diabetes | COVID-19 infection | Asthma | Chronic Kidney Disease (CKD) | Secondary pulmonary hypertension | VPC Field Completion |
|---|---|---|---|---|---|---|---|---|---|---|---|---|---|---|
| Algorithm ID | Y | 1 | 1 | 1 | 1 | 1 | **1** | Y | 1 | 1 | 1 | 1 | 1 | **1** |
| Title | Y | 1 | 1 | 1 | 1 | 1 | **1** | Y | 1 | 1 | 1 | 1 | 1 | **0.9** |
| Definition | Y | 0 | 1 | 1 | 1 | 1 | **0.8** | Y | 0 | 1 | 1 | 1 | 1 | **0.9** |
| Phenotype type | Y | 1 | 1 | 1 | 1 | 1 | **1** | Y | 1 | 1 | 1 | 1 | 1 | **1** |
| Primary use population | Y | 1 | 1 | 1 | 1 | 1 | **1** | Y | 1 | 1 | 1 | 1 | 1 | **1** |
| Primary use population subcategory | Y | 1 | 1 | 1 | 1 | 1 | **1** | Y | 1 | 1 | 1 | 1 | 1 | **1** |
| Authors | Y | 1 | 1 | 1 | 1 | 1 | **1** | Y | 1 | 1 | 1 | 1 | 1 | **0.6** |
| Corresponding contact | Y | 0 | 0 | 0 | 0 | 1 | **0.2** | Y | 0 | 0 | 0 | 0 | 1 | **0.6** |
| Created date | Y | 1 | 1 | 1 | 1 | 1 | **1** | Y | 1 | 1 | 1 | 1 | 1 | **1** |
| Last updated | Y | 1 | 1 | 1 | 1 | 1 | **1** | Y | 1 | 1 | 1 | 1 | 1 | **0.5** |
| License | N | *N/A* | *N/A* | *N/A* | *N/A* | *N/A* | **N/A** | Y | 0 | 0 | 0 | 0 | 0 | **0** |
| Primary use case | N | *N/A* | *N/A* | *N/A* | *N/A* | *N/A* | **N/A** | Y | 0 | 0 | 0 | 0 | 0 | **0.5** |
| URL for resource | Y | 1 | 1 | 1 | 1 | 1 | **1** | Y | 1 | 1 | 1 | 1 | 1 | **0.9** |
| Publications | Y | 1 | 0 | 1 | 1 | 1 | **0.8** | Y | 1 | 0 | 1 | 1 | 1 | **0.8** |
| Source data | Y | 1 | 1 | 1 | 1 | 1 | **1** | Y | 1 | 1 | 1 | 1 | 0 | **0.9** |
| Data source terminologies | Y | 1 | 1 | 1 | 1 | 1 | **1** | Y | 1 | 1 | 1 | 1 | 1 | **1** |
| Approach | Y | N/A | *N/A* | *N/A* | *N/A* | *N/A* | **N/A** | Y | 1 | 1 | 1 | 1 | 1 | **0.7** |
| Algorithm description | Y | 0 | 0 | 0 | 0 | 1 | **0.2** | Y | 0 | 0 | 0 | 1 | 1 | **0.4** |
| URL for source code | Y | 0 | 1 | 1 | 0 | 0 | **0.4** | Y | 0 | 1 | 1 | 0 | 0 | **0.2** |
| Validation population | Y | 0 | 0 | 0 | 0 | 1 | **0.2** | Y | 0 | 0 | 0 | 0 | 0 | **0** |
| Reference standard | N | *N/A* | *N/A* | *N/A* | *N/A* | *N/A* | **N/A** | Y | 0 | 0 | 0 | 0 | 0 | **0** |
| Guideline | N | *N/A* | *N/A* | *N/A* | *N/A* | *N/A* | **N/A** | Y | 0 | 0 | 0 | 0 | 0 | **0** |
| Metrics | N | *N/A* | *N/A* | *N/A* | *N/A* | *N/A* | **N/A** | Y | 0 | 0 | 0 | 0 | 0 | **0** |
| Fields populated of the 23 framework fields | | 12 | 13 | 14 | 13 | 16 | | | 13 | 14 | 15 | 15 | 15 | |
| Completion, all 23 fields | | 52% | 57% | 61% | 57% | 70% | | | 57% | 61% | 65% | 65% | 65% | |
| Completion, schema-supported fields | | 67% | 72% | 78% | 72% | 89% | | | 57% | 61% | 65% | 65% | 65% | |
| Library mean, all 23 fields | 59% | | | | | | | 63% | | | | | | |
| Library mean, schema-supported fields only | 76% | | | | | | | 63% | | | | | | |
