## Supplemental table 7 for "OPEN: A Community-Informed Metadata Framework for Interoperable Computable Phenotypes"

**Supplementary Table 7: HDR UK to VPC Library Mapping**

| HDR UK Field | HDR UK Value | Field Complete (0/1) | Metadata Framework Field | VPC Library Field | VPC Library Value | Field Complete (0/1) | Note |
|---|---|---|---|---|---|---|---|
| phenotype_id, phenotype_version_ | PH1741/3673 | 1 | Algorithm ID | | 110 | 1 | Auto genera |
| name | Diabetes | 1 | Title | Phenotype Name | Diabetes | 1 | |
| Definition | | 0 | Definition | Phenotype Description | | 0 | |
| Phenotype Type | Disease or syndrome | 1 | Phenotype type | Phenotype Type | Disease or syndrome | 1 | |
| data_sources[].name ? | Trusted Research Environments for ( | 1 | Primary use population | Primary Site | Trusted Research Environments | 1 | |
| Sex; Valid event date range; | Sex: Both; Valid event date range: n | 1 | Primary use population subcategory | Sex; Age Group; Age Range | Sex: Both; | 1 | |
| author | Ip, Torabi, Denaxas, Akbari ... Walke | 1 | Authors | Authors | Ip, Torabi, Denaxas, Akbari ... W | 1 | |
| corresp_author, corresp_email | | 0 | Corresponding contact | Contact | | 0 | |
| created | 4/7/2025 | 1 | Created date | Authored on | 4/7/2025 | 1 | |
| updated | 4/7/2025 | 1 | Last updated | Last Updated | 4/7/2025 | 1 | Automatica |
| *N/A* | *N/A* | *N/A* | License | License | | 0 | Not contain |
| *N/A* | *N/A* | *N/A* | Primary use case | Primary Use Case | | 0 | |
| doi | https://doi.org/10.48533/tm7r-2b39 | 1 | URL for resource | Resource URL | https://doi.org/10.48533/tm7r- | 1 | |
| publication | https://doi.org/10.1101/2022.03.06. | 1 | Publications | Publications | https://doi.org/10.1101/2022.0 | 1 | |
| Data Sources | Diagnoses: Yes; Procedures: Yes | 1 | Source data | Data Source | Diagnoses: Yes; Procedures: Yes | 1 | |
| Coding System | SNOMED CT codes, ICD10 codes | 1 | Data source terminologies | Terminologies Used | SNOMED CT, ICD-10 | 1 | |
| template.name | BHF Data Science Centre Phenotype | *N/A* | Approach | Algorithm Type | BHF Data Science Centre Pheno | 1 | |
| implementation | *Null* | 0 | Algorithm description | Algorithm Description | | 0 | |
| phenoflowid | *Null* | 0 | URL for source code | Source Code URL | | 0 | |
| validation | *Null* | 0 | Validation population | Validation Population | | 0 | |
| *N/A* | *N/A* | *N/A* | Reference standard | Reference Standard | | 0 | |
| *N/A* | *N/A* | *N/A* | Guideline | Guideline | | 0 | |
| *N/A* | *N/A* | *N/A* | Metrics | Performance Metrics | Sensitivity; Specificity; PPV; NPV; AUF | 0 | |

| HDR UK Field | HDR UK Value | Field Complete (0/1) | Metadata Framework Field | VPC Library Field | VPC Library Value | Field Complete (0/1) | Note |
|---|---|---|---|---|---|---|---|
| phenotype_id, phenotype_version_ | PH1 / 2 | 1 | Algorithm ID | | 111 | 1 | Auto genera |
| name | COVID-19 infection | 1 | Title | Phenotype Name | COVID-19 infection | 1 | |
| Definition | We ascertained people with a confir | 1 | Definition | Phenotype Description | We ascertained people with a c | 1 | |
| Phenotype Type | Disease or syndrome | 1 | Phenotype type | Phenotype Type | Disease or syndrome | 1 | |
| data_sources[].name ? | GPES Data for Pandemic Planning ar | 1 | Primary use population | Primary Site | GPES Data for Pandemic Plannir | 1 | |
| Sex; Valid event date range; | Sex: Both; Valid Event Date Range: 0 | 1 | Primary use population subcategory | Sex; Age Group; Age Range | Sex: Both; | 1 | |
| author | BHF CVD COVID UK Consortium | 1 | Authors | Authors | BHF CVD COVID UK Consortium | 1 | |
| corresp_author, corresp_email | *Null* | 0 | Corresponding contact | Contact | | 0 | |
| created | 10/6/2021 | 1 | Created date | Authored on | 10/6/2021 | 1 | |
| updated | 10/6/2021 | 1 | Last updated | Last Updated | 1-Sep-26 | 1 | |
| *N/A* | *N/A* | *N/A* | License | License | | 0 | Not contain |
| *N/A* | *N/A* | *N/A* | Primary use case | Primary Use Case | | 0 | |
| doi | https://doi.org/10.48533/wjmc-hh1 | 1 | URL for resource | Resource URL | https://doi.org/10.48533/wjmc- | 1 | |
| publication | *Null* | 0 | Publications | Publications | | 0 | |
| Data Sources | https://healthdatagateway.org/en/c | 1 | Source data | Data Source | https://healthdatagateway.org/ | 1 | |
| Coding System | ICD10 codes, SNOMED CT codes | 1 | Data source terminologies | Terminologies Used | ICD10 codes, SNOMED CT code: | 1 | |

| | | | | | | |
|---|---|---|---|---|---|---|
| template.name | BHF Data Science Centre Phenotype | *N/A* | Approach | Algorithm Type | BHF Data Science Centre Pheno | 1 |
| implementation | *Null* | 0 | Algorithm description | Algorithm Description | | 0 |
| phenoflowid | https://kclhi.org/phenoflow/workflc | 1 | URL for source code | Source Code URL | https://kclhi.org/phenoflow/wc | 1 |
| validation | *Null* | 0 | Validation population | Validation Population | | 0 |
| *N/A* | *N/A* | *N/A* | Reference standard | Reference Standard | | 0 |
| *N/A* | *N/A* | *N/A* | Guideline | Guideline | | 0 |
| *N/A* | *N/A* | *N/A* | Metrics | Performance Metrics | Sensitivity; Specificity; PPV; NPV; AUF | 0 |

| **HDR UK Field** | **HDR UK Value** | **Field Complete (0/1)** | **Metadata Framework Field** | **VPC Library Field** | **VPC Library Value** | **Field Complete (0/1)** | **Note** |
|---|---|---|---|---|---|---|---|
| phenotype_id, phenotype_version_i | PH12 / 24 | 1 | Algorithm ID | | 112 | 1 | Auto genera |
| name | Asthma | 1 | Title | Phenotype Name | Asthma HDRUK | 1 | |
| Definition | Captures asthma ever, not current a | 1 | Definition | Phenotype Description | Captures asthma ever, not curre | 1 | |
| Phenotype Type | Disease or syndrome | 1 | Phenotype type | Phenotype Type | Disease or syndrome | 1 | |
| data_sources[].name ? | CPRD GOLD; CPRD Aurum; HES APC | 1 | Primary use population | Primary Site | CPRD GOLD; CPRD Aurum; HES | 1 | |
| Sex; Valid event date range; | Sex: Both; Valid Event Date Range: 0 | 1 | Primary use population subcategory | Sex; Age Group; Age Range | Sex: Both; | 1 | |
| author | Eleanor L Axson, Jennifer K Quint | 1 | Authors | Authors | Eleanor L Axson, Jennifer K Quir | 1 | |
| corresp_author, corresp_email | ieuan.scanlon | 0 | Corresponding contact | Contact | | 0 | |
| created | 10/6/2021 | 1 | Created date | Authored on | 10/6/2021 | 1 | |
| updated | 10/6/2021 | 1 | Last updated | Last Updated | 10/6/2021 | 1 | |
| *N/A* | *N/A* | *N/A* | License | License | | 0 | Not contain |
| *N/A* | *N/A* | *N/A* | Primary use case | Primary Use Case | | 0 | |
| doi | https://doi.org/10.48533/aw5j-jh79 | 1 | URL for resource | Resource URL | https://doi.org/10.48533/aw5j- | 1 | |
| publication | "F. Nissen, Douglas, I. J., Mullerova, | 1 | Publications | Publications | "F. Nissen, Douglas, I. J., Muller | 1 | |
| Data Sources | Diagnoses: Yes; Mortality: Yes; Surve | 1 | Source data | Data Source | CPRD GOLD; CPRD Aurum; HES | 1 | |
| Coding System | ICD10, Read v2, SNOMED CT, UKBiol | 1 | Data source terminologies | Terminologies Used | ICD10, Read v2, SNOMED CT, UI | 1 | |
| template.name | Clinical-Coded Phenotype | *N/A* | Approach | Algorithm Type | Clinical-Coded Phenotype | 1 | |
| implementation | *Null* | 0 | Algorithm description | Algorithm Description | | 0 | |
| phenoflowid | https://kclhi.org/phenoflow/workflc | 1 | URL for source code | Source Code URL | https://kclhi.org/phenoflow/wc | 1 | |
| validation | *Null* | 0 | Validation population | Validation Population | | 0 | |
| *N/A* | *N/A* | *N/A* | Reference standard | Reference Standard | | 0 | |
| *N/A* | *N/A* | *N/A* | Guideline | Guideline | | 0 | |
| *N/A* | *N/A* | *N/A* | Metrics | Performance Metrics | Sensitivity; Specificity; PPV; NPV; AUF | 0 | |

| **HDR UK Field** | **HDR UK Value** | **Field Complete (0/1)** | **Metadata Framework Field** | **VPC Library Field** | **VPC Library Value** | **Field Complete (0/1)** | **Note** |
|---|---|---|---|---|---|---|---|
| phenotype_id, phenotype_version_i | PH22 / 44 | 1 | Algorithm ID | | 113 | 1 | Auto genera |
| name | Chronic Kidney Disease (CKD) | 1 | Title | Phenotype Name | Chronic Kidney Disease (CKD) | 1 | |
| Definition | The data (demographics, emergency | 1 | Definition | Phenotype Description | The data (demographics, emerg | 1 | |
| Phenotype Type | Disease or syndrome | 1 | Phenotype type | Phenotype Type | Disease or syndrome | 1 | |
| data_sources[].name ? | King's College Hospital NHS Foundat | 1 | Primary use population | Primary Site | King's College Hospital NHS Fou | 1 | |
| Sex; Valid event date range; | Sex: Both; Valid Event Date Range: 0 | 1 | Primary use population subcategory | Sex; Age Group; Age Range | Sex: Both; | 1 | |

| | | | | | | | |
|---|---|---|---|---|---|---|---|
| author | Carr E, Bendayan R, Bean D, O'Gallag | 1 | Authors | Authors | Carr E, Bendayan R, Bean D, O'G | 1 | |
| corresp_author | *Null* | 0 | Corresponding contact | Contact | | 0 | |
| created | 10/6/2021 | 1 | Created date | Authored on | 10/6/2021 | 1 | |
| updated | 10/6/2021 | 1 | Last updated | Last Updated | 1-Sep-26 | 1 | |
| *N/A* | *N/A* | *N/A* | License | License | | 0 | Not contain |
| *N/A* | *N/A* | *N/A* | Primary use case | Primary Use Case | | 0 | |
| doi | https://doi.org/10.48533/pmj9-m27 | 1 | URL for resource | Resource URL | https://doi.org/10.48533/pmj9- | 1 | |
| publication | https://doi.org/10.1101/2020.04.24. | 1 | Publications | Publications | https://doi.org/10.1101/2020.0 | 1 | |
| Data Sources | Diagnoses: Yes | 1 | Source data | Data Source | King's College Hospital NHS Fou | 1 | |
| Coding System | SNOMED CT codes | 1 | Data source terminologies | Terminologies Used | SNOMED CT codes | 1 | |
| template.name | Clinical-Coded Phenotype | *N/A* | Approach | Algorithm Type | Rule Based | 1 | |
| implementation | *Null* | 0 | Algorithm description | Algorithm Description | Clinical-Coded Phenotype | 1 | |
| phenoflowid | *Null* | 0 | URL for source code | Source Code URL | | 0 | |
| validation | *Null* | 0 | Validation population | Validation Population | | 0 | |
| *N/A* | *N/A* | *N/A* | Reference standard | Reference Standard | | 0 | |
| *N/A* | *N/A* | *N/A* | Guideline | Guideline | | 0 | |
| *N/A* | *N/A* | *N/A* | Metrics | Performance Metrics | Sensitivity; Specificity; PPV; NPV; AUF | 0 | |

| **HDR UK Field** | **HDR UK Value** | **Field Complete (0/1)** | **Metadata Framework Field** | **VPC Library Field** | **VPC Library Value** | **Field Complete (0/1)** | **Note** |
|---|---|---|---|---|---|---|---|
| phenotype_id, phenotype_version_ | PH4349 / 9567 | 1 | Algorithm ID | | 114 | 1 | Auto genera |
| name | Secondary pulmonary hypertension | 1 | Title | Phenotype Name | Secondary pulmonary hyperten | 1 | |
| Definition | Raised pulmonary vascular pressure | 1 | Definition | Phenotype Description | Raised pulmonary vascular pres | 1 | |
| Phenotype Type | Disease or syndrome | 1 | Phenotype type | Phenotype Type | Disease or syndrome | 1 | |
| data_sources[].name ? | UK Biobank; Patient Episode Dataset | 1 | Primary use population | Primary Site | UK Biobank; Patient Episode Da | 1 | |
| Sex; Valid event date range; | Sex: Both; Valid event date range: n | 1 | Primary use population subcategory | Sex; Age Group; Age Range | Sex: Both; | 1 | |
| author | Torralbo A; Davitte JM; Croteau-Cho | 1 | Authors | Authors | Torralbo A; Davitte JM; Croteau | 1 | |
| corresp_author | Ana Torralbo, | 1 | Corresponding contact | Contact | | 1 | |
| created | 6/8/2026 | 1 | Created date | Authored on | 6/8/2026 | 1 | |
| updated | 6/8/2026 | 1 | Last updated | Last Updated | 6/8/2026 | 1 | |
| *N/A* | *N/A* | *N/A* | License | License | | 0 | Not contain |
| *N/A* | *N/A* | *N/A* | Primary use case | Primary Use Case | | 0 | |
| doi | https://doi.org/10.48533/d8qd-2p7 | 1 | URL for resource | Resource URL | https://doi.org/10.48533/d8qd- | 1 | |
| publication | https://doi.org/10.1038/s41598-025 | 1 | Publications | Publications | https://doi.org/10.1038/s41598 | 1 | |
| Data Sources | Diagnoses: Yes; Mortality: Yes | 1 | Source data | Data Source | | 0 | |
| Coding System | ICD10 codes, Read codes v3 | 1 | Data source terminologies | Terminologies Used | ICD10 codes, Read codes v3 | 1 | |
| template.name | BHF Data Science Centre Phenotype | *N/A* | Approach | Algorithm Type | Rule based | 1 | |
| implementation | The earliest record across all source | 1 | Algorithm description | Algorithm Description | The earliest record across all so | 1 | |
| phenoflowid | *Null* | 0 | URL for source code | Source Code URL | | 0 | |
| validation | Multi-layered validation: data sourc | 1 | Validation population | Validation Population | | 0 | |
| *N/A* | *N/A* | *N/A* | Reference standard | Reference Standard | | 0 | |
| *N/A* | *N/A* | *N/A* | Guideline | Guideline | | 0 | |

| | | | | | | | |
|---|---|---|---|---|---|---|---|
| *N/A* | *N/A* | *N/A* | Metrics | Performance Metrics | Sensitivity; Specificity; PPV; NPV; AUF | 0 | FPR is missi |
